# Detection of the omicron variant virus with the Abbott BinaxNow SARS-CoV-2 Rapid Antigen Assay

**DOI:** 10.1101/2021.12.22.21268219

**Authors:** James Regan, James P. Flynn, Manish C. Choudhary, Rockib Uddin, Jacob Lemieux, Julie Boucau, Roby P. Bhattacharyya, Amy K. Barczak, Jonathan Z. Li, Mark J. Siedner

## Abstract

The US Centers for Disease Control and Prevention recommends rapid testing for SARS-CoV-2 infection as a key element of epidemic control. The Abbott BinaxNow is in widespread use in the United States for self-testing and as part of public health screening campaigns, but has not been evaluated for use with the omicron variant of SARS-CoV-2. We recruited individuals testing positive for COVID-19 PCR at an academic medical center. Anterior nasal swabs were stored in viral transport media and evaluated by viral load quantification and whole genome sequencing. We created serial dilutions from 2.5×10^3^-2.5×10^5^ viral copies/specimen for two delta and omicron specimens, respectively, and tested each with the BinaxNow assay per manufacturer instructions. Results were interpreted by three readers, blinded to the specimen variant and concentration. All omicron and delta specimens with concentrations of 100,000 copies/swab or greater were positive by the BinaxNow Assay, a concentration similar to previously reported limits of detection for this assay. Assay sensitivity diminished below that. This study demonstrates that Omicron variant SARS-CoV-2 infections are detected by the BinaxNow rapid antigen assay. Additional laboratory and clinical validation assessments are needed to better determine their limits of detection and performance in real-world settings.

## Background

The US Centers for Disease Control and Prevention recommends rapid testing for SARS-CoV-2 infection as a key element of epidemic control (1). One such assay, the Abbott BinaxNow, has been recommended for in-home testing and implemented in public health screening campaigns (2–6). The diagnostic validity of the assay has been demonstrated in laboratory-based and clinical settings, with a sensitivity ranging from approximately 50-90%, depending on disease stage and degree of symptom, and a specificity >99%, compared to laboratory-based PCR assays (3,4,7,8).

Notably, test performance of chromatographic immunoassays such as the BinaxNow is dependent on specific viral antigens and may be impacted by changes in viral protein structure. In November 2021, the omicron variant of SARS-CoV-2 was first detected in Southern Africa, and quickly disseminated globally (9). The initially identified lineage of the omicron variant contains more than 50 mutations and deletions in comparison to ancestral lineages, including four mutations in the nucleocapsid gene, which is the target for the BinaxNow assay. Abbott, the manufacturer of the assay has reported in a press release that the assay is predicted to detect the omicron variant of SARS-CoV-2 infection (10). However, there are no published data on the validity of the assay with clinical specimens.

## Methods

We conducted a laboratory-based validation of the BinaxNow assay with anterior nasal (AN) swab specimens from participants in a study of COVID-19 virology (11). We recruited individuals testing positive for COVID-19 PCR at an academic medical center. Positive AN swabs were stored in viral transport media and evaluated by viral load quantification and whole genome sequencing (12). We selected two omicron variant specimens and two delta variant specimens and produced serial dilutions to create swabs of 2.5×10^5^, 1.0×10^5^, 2.5×10^4^, and 2.5×10^3^ viral copies with each specimen. These dilutions were chosen to create concentrations both above and below previously reported limits of detection of the assay in a laboratory-based evaluation (8). To do so, we initially created viral concentrations of 5.0×10^4^, 5.0×10^5^, 2.0×10^6^, 5.0×10^6^ copies/mL in phosphate buffer saline, then immersed swabs from the BinaxNow kits into 50µL of each dilution until the material was fully absorbed, as previously described (8). An additional specimen with only phosphate-buffered saline was used as a negative control. The swabs were then tested according to the manufacturer’s instructions and results interpreted by three readers, blinded to the specimen variant and concentration, as positive, negative, or discordant (if not all three readers agreed). Study procedures were approved by the institutional review board of Mass General Brigham and participants gave consent for specimen testing for research purposes.

## Results

The four specimens with concentrations of 100,000 copies/swab or greater were positive with both the delta and omicron variant specimens (Figure 1). Assay sensitivity was diminished below that, with positive results in 1/4 omicron specimens and 1/3 of delta specimens (with the fourth delta specimen resulting in discordant reads between reviewers). All specimens with 2,500 copies/swab were interpreted as negative.

**Figure 1.**
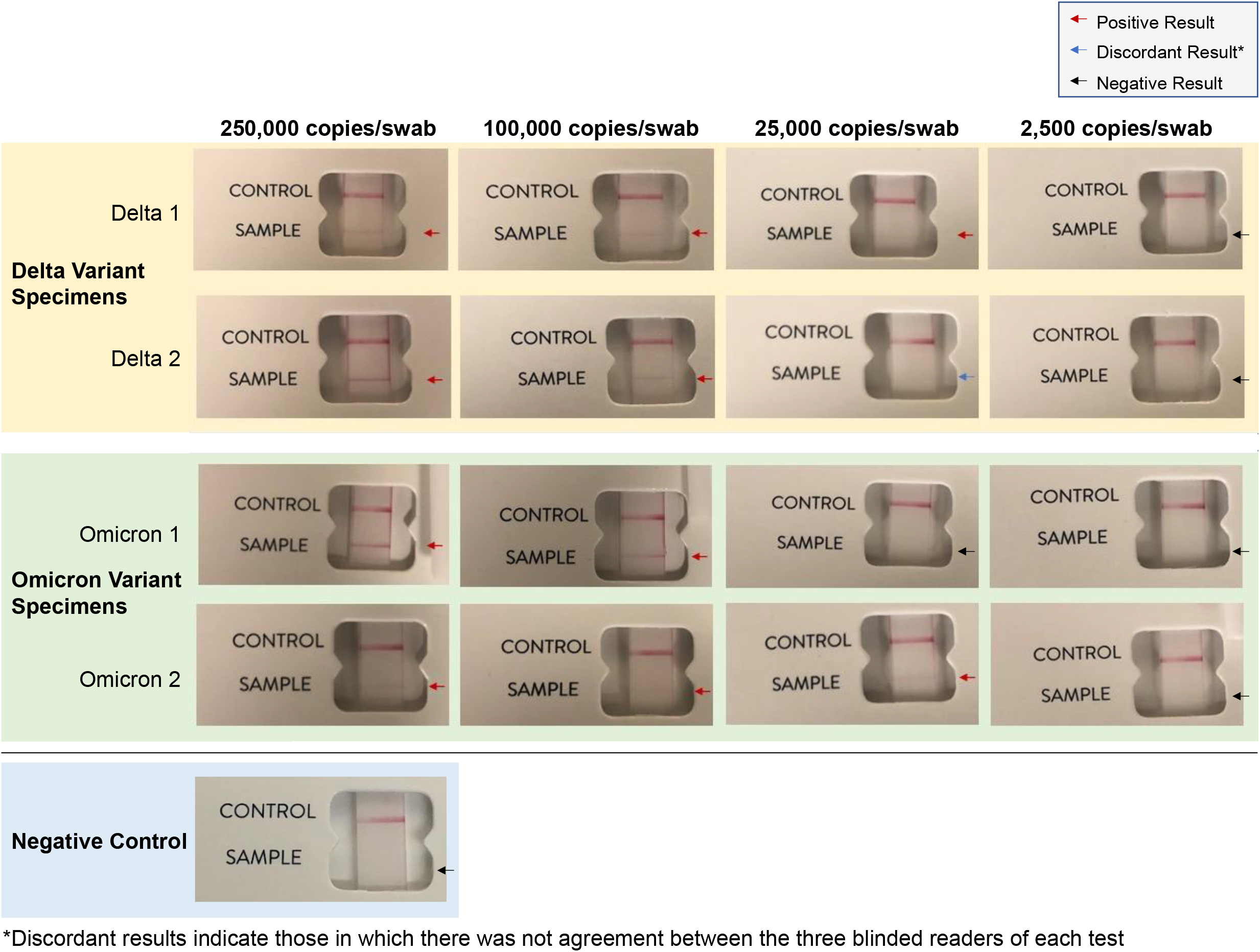
Results of a laboratory-based analytic validation of the BinaxNow SARS-CoV-2 rapid antigen assay across a range of concentrations with both delta and omicron variant viruses

## Conclusions

Omicron variant SARS-CoV-2 infections were detected in a laboratory-based assessment of the BinaxNow rapid antigen assay. This is the first report to our knowledge of this assay being formally assessed with the omicron variant. Although our study does not intend to identify a limit of detection for this assay, our results are qualitatively similar to previously published results suggesting the BinaxNow assay has a limit of detection of approximately 2.0×10^4^ - 7.0×10^4^ viral copies/swab (8). These data offer proof of concept that the BinaxNow rapid antigen assay can detect the Omicron variant SARS-CoV-2 infections. Future work should more thoroughly assess the diagnostic validity and range of detection of the assay for this novel variant, as well as its performance in clinical, public health and self-testing scenarios.

## Data Availability

All data produced in the present study are available upon reasonable request to the authors

## References

1. Interim Guidance for Antigen Testing for SARS-CoV-2. United States Centers for Disease Control and Prevention. [Internet]. 2020 [cited 2021 Dec 20]. Available from: https://www.cdc.gov/coronavirus/2019-ncov/lab/resources/antigen-tests-guidelines.html

2. Prince-Guerra JL, Almendares O, Nolen LD, Gunn JKL, Dale AP, Buono SA, et al. Evaluation of Abbott BinaxNOW Rapid Antigen Test for SARS-CoV-2 Infection at Two Community-Based Testing Sites — Pima County, Arizona, November 3–17, 2020. MMWR Morb Mortal Wkly Rep. 2021 Jan 22;70(3):100–5.

3. Okoye NC, Barker AP, Curtis K, Orlandi RR, Snavely EA, Wright C, et al. Performance Characteristics of BinaxNOW COVID-19 Antigen Card for Screening Asymptomatic Individuals in a University Setting. Journal of Clinical Microbiology [Internet]. 2021 Jan 28[cited 2021 Dec 20]; Available from: https://journals.asm.org/doi/abs/10.1128/JCM.03282-20

4. Almendares O, Prince-Guerra JL, Nolen LD, Gunn JKL, Dale AP, Buono SA, et al. Performance characteristics of the Abbott BinaxNOW SARS-CoV-2 antigen test in comparison to real-time RT-PCR and viral culture in community testing sites during November 2020. Journal of Clinical Microbiology [Internet]. 2021 Oct 27[cited 2021 Dec 20]; Available from: https://journals.asm.org/doi/abs/10.1128/JCM.01742-21

5. Tinker SC, Szablewski CM, Litvintseva AP, Drenzek C, Voccio GE, Hunter MA, et al. Point-of-Care Antigen Test for SARS-CoV-2 in Asymptomatic College Students. Emerg Infect Dis. 2021 Oct;27(10):2662–5.

6. Rubin R. COVID-19 Testing Moves Out of the Clinic and Into the Home. JAMA. 2021 Oct 12;326(14):1362–4.

7. Siddiqui ZK, Chaudhary M, Robinson ML, McCall AB, Peralta R, Esteve R, et al. Implementation and Accuracy of BinaxNOW Rapid Antigen COVID-19 Test in Asymptomatic and Symptomatic Populations in a High-Volume Self-Referred Testing Site. Microbiology Spectrum [Internet]. 2021 Dec 1[cited 2021 Dec 20]; Available from: https://journals.asm.org/doi/abs/10.1128/Spectrum.01008-21

8. Perchetti GA, Huang M-L, Mills MG, Jerome KR, Greninger AL. Analytical Sensitivity of the Abbott BinaxNOW COVID-19 Ag Card. Journal of Clinical Microbiology [Internet]. 2020 Dec 11[cited 2021 Dec 20]; Available from: https://journals.asm.org/doi/abs/10.1128/JCM.02880-20

9. Karim SSA, Karim QA. Omicron SARS-CoV-2 variant: a new chapter in the COVID-19 pandemic. The Lancet. 2021 Dec 11;398(10317):2126–8.

10. Evaluating Omicron and Other COVID Variants to Ensure Test Effectiveness. Abbott. Available at: https://www.abbott.com/corpnewsroom/diagnostics-testing/monitoring-covid-variants-to-ensure-test-effectiveness.html. Accessed 20 Dec 2021.

11. Siedner MJ, Boucau J, Gilbert RF, Uddin R, Luu J, Haneuse S, et al. Duration of viral shedding and culture positivity with post-vaccination SARS-CoV-2 delta variant infections. JCI Insight. 2021 Dec 6;e155483.

12. The Massachusetts Consortium for Pathogen Readiness, Fajnzylber J, Regan J, Coxen K, Corry H, Wong C, et al. SARS-CoV-2 viral load is associated with increased disease severity and mortality. Nat Commun. 2020 Dec;11(1):5493.

